# Short-term ambient heat exposure and low APGAR score in newborns: A time-stratified case-crossover analysis in São Paulo state, Brazil (2013-2019)

**DOI:** 10.1101/2025.04.08.25325468

**Authors:** Michelle Del Carretto, Audrey Godin, Danielson Neves, Enny S. Paixao, Kai Wan, Julia Pescarini, Andrêa Ferreira, Taisa R. Cortes, Liam Smeeth, Mauricio L. Barreto, Elizabeth B. Brickley, Chérie Part

## Abstract

Exposure to high ambient temperatures near the time of delivery has been associated with adverse birth outcomes, but studies examining the impact on immediate newborn health remain limited. We used a time-stratified case-crossover design combined with a distributed lag nonlinear model to evaluate the short-term effects of ambient heat (0-1 day lag) on low 5-minute APGAR score (≤7; sub-categories: 6-7, 3-5, 0-2). Cases of low APGAR score among low-risk births (*n*=34,980) in São Paulo state (274 municipalities), 2013-2019, were extracted from Brazil’s Live Birth Information System (*Sistema de Informações Sobre Nascidos Vivos*). Municipality-level daily mean temperatures were constructed from ERA5-Land reanalysis data and linked with case and control days by date and municipality of delivery. Models were adjusted for relative humidity and stratified by maternal age, race/ethnicity, education, parity, timing of prenatal care initiation, infant sex, municipality-level deprivation, and Köppen climate zone. Overall, exposure to high (95^th^ percentile: 26.1°C) versus moderate (50^th^ percentile: 20.9°C) temperature 0-1 days before delivery was associated with 8% higher odds (OR: 1.08, 95% CI: 1.02-1.14) of low APGAR score (≤7). In stratified analyses, heat-associated risks were elevated among infants born to women with <12 years of schooling (1.10, 1.03-1.17) and/or self-identifying as Brown/*Parda* (1.10, 1.01-1.20). Associations were primarily driven by same-day (lag 0) exposure and were only observed in newborns with moderately low APGAR scores (6-7). Acute exposure to ambient heat may adversely impact newborns’ immediate health in low-risk live-births, highlighting the need for heat mitigation measures near the time of delivery.

## Introduction

Rising global temperatures pose a substantial threat to maternal and newborn health (1). Exposure to ambient heat during pregnancy increases the risk of maternal complications (2) and adverse birth outcomes, such as preterm birth, stillbirth, and low birth weight (3). Studies focusing on neonatal outcomes remain scarce. There is evidence for an increased risk of neonatal hospital admissions during heatwaves in Brazil and India (4,5), however these studies have limited ability to determine critical windows of susceptibility. Neonatal intensive care admissions include in-hospital transfers immediately post-birth as well as admissions up to 28 days of life. Even when examining the effects of same-day exposure among term births, studies have yet to determine the stage at which exposure to ambient heat poses the greatest risk (4).

The APGAR score is used worldwide to measure newborn health and response to resuscitation interventions at 1-, 5- and 10-minutes of life (6). A low score indicates poor immediate health condition, and has been associated with long-term health concerns, such as cerebral palsy and epilepsy (7). Prematurity and low birth weight are risk factors for low APGAR scores. However, even among term, optimal birth weight newborns, this measure is particularly sensitive to characteristics of the labour and delivery, including prolonged labour and surgical (caesarean) birth (8,9).

There is evidence that low APGAR scores are associated with exposure to high ambient temperatures during the third trimester of pregnancy (10–13). However, previous studies often averaged temperature exposures over broad windows of susceptibility, such as months or trimesters. We hypothesise that acute exposure to ambient heat, during labour and delivery, influences newborns’ immediate health through several interconnected mechanisms (Fig 1). Circulatory system changes (e.g. dehydration, vasodilation) can reduce uterine-placental blood flow, hindering blood-oxygen exchange with the foetus (14) or triggering maternal cardiovascular events (15). Inflammatory pathways may cause Premature Rupture of Membranes when combined with prior infection (16). Discomfort and heat stress may cause fatigue and prolonged labour (17). These pathways may also increase the need for surgical intervention during delivery, contributing to foetal distress (18). Heat effects on maternal and newborn health may also interact with aspects of delivery care (e.g. healthcare workforce strain and care quality), living conditions (e.g., limited access to air conditioning) and environment (e.g., air pollution), all of which are inherently interlinked with the social determinants of health (19,20).

**Fig 1.**
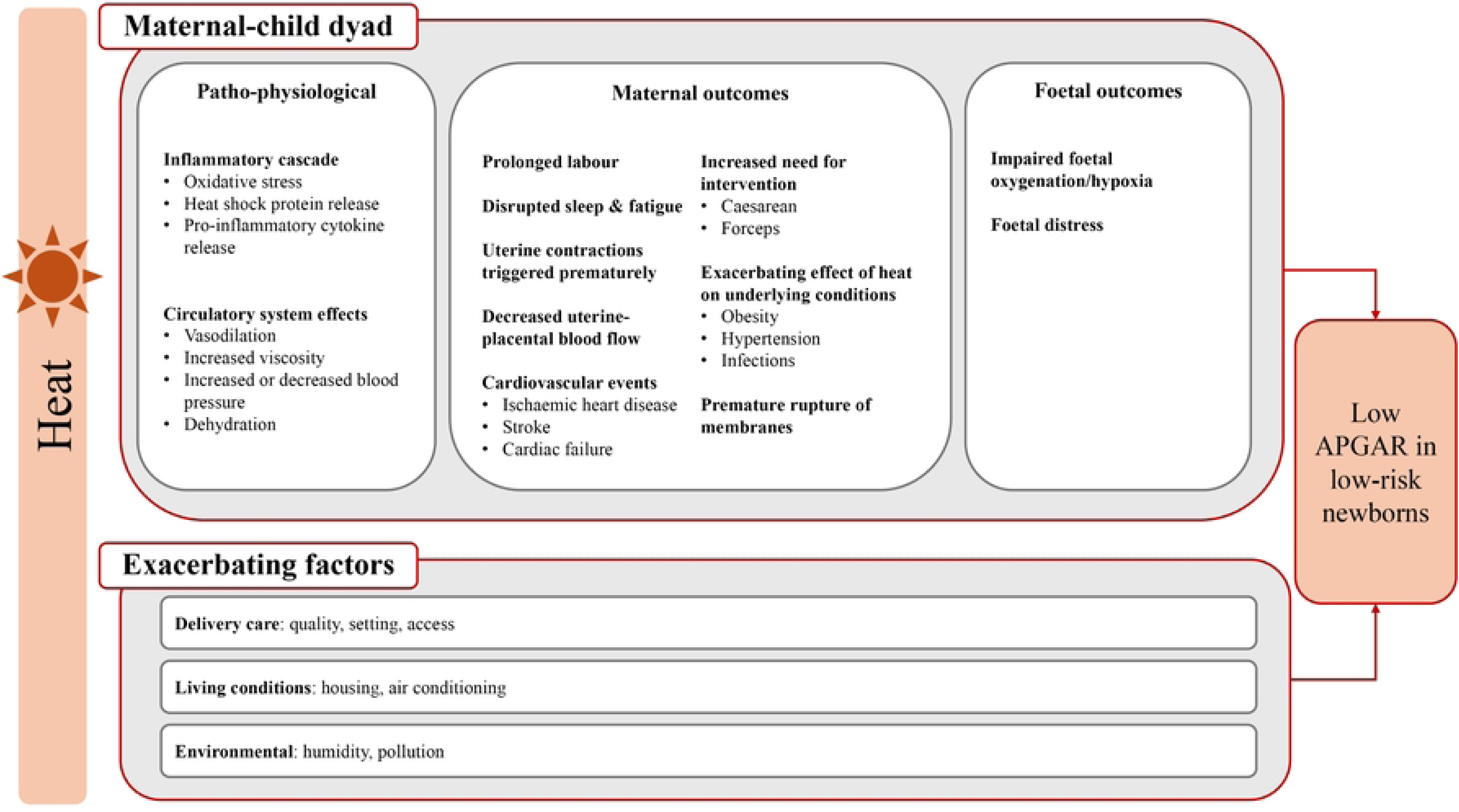
Hypothesised mechanisms through which short-term ambient heat exposure increases the risk of low APGAR-5’ scores.

Using a time-stratified case-crossover design, we evaluated the short-term association between daily mean temperature and the odds of low APGAR score at 5-minutes of life (APGAR-5’) in São Paulo state, Brazil. Cases were restricted to low-risk live-births (defined here as singleton, term births, with optimal birth weight, cephalic presentation, and absence of congenital anomalies) to remove competing biological pathways. São Paulo state is situated in the Southeast region of Brazil, where the last decade has seen rising temperatures and more frequent, intense heatwaves (21). Given the unequal burden of heat-health outcomes according to education level and race/ethnicity in Brazil (22,23), subgroup analyses of socio-economic relevance were conducted in addition to those of clinical significance to identify high-risk groups for targeted intervention.

## Methods

### Study setting

São Paulo state is highly urbanised, with a total population of more than 44 million (24). As defined by Köppen climate designations, the state is characterised by temperate (C) and tropical (A) climates, with a majority of the population living in humid-subtropical, oceanic areas (Cfa, Cfb - hot, humid or mild summers and mild winters, Fig S1) (25). Extremely hot summers of 2013/14 and 2014/15 highlight the state’s vulnerability to climate change (21). Approximately 99% of births in Brazil occur in hospitals (26). Despite Brazil’s universal healthcare system providing free, essential maternity services, São Paulo state suffers from inequalities in access for certain ethno-racial and socio-economic groups (27,28).

### Ethics statement

As the study analyzed de-identified publicly available data, ethical approval was not required in accordance with Resolução N° 510 (7 April 2016) of the Brazilian Ethics System (Sistema CEP-CONEP) and confirmed with the London School of Hygiene and Tropical Medicine Research Ethics Committee (Reference 30472 and 30657).

### Data

#### Birth data and outcome assessment

De-identified, individual-level data on live births were downloaded from the Brazilian Ministry of Health’s Unified Health System data registry (*Departamento de Informática do Sistema Único de Saúde*, DATASUS) on 28 May 2024 using the Live Births Information System (*Sistema de Informações Sobre Nascidos Vivos*, SINASC). SINASC contains information on the pregnant person (e.g. age, municipality of delivery), the pregnancy (e.g. completed weeks’ gestation) and the newborn (e.g. APGAR scores, birth weight).

Between 01 January 2013 and 31 December 2019, 4,166,174 live, singleton births were registered in São Paulo state. Births occurring outside the state (N= 4,575; 0.11%) and those with missing 5-minute APGAR score (N=19,404; 0.47%) were removed. Data were restricted to newborns from low-risk births by excluding infants with: (i) birth weight <2500 or >4000 grams (N=499,493; 11.99%), (ii) gestational age <37 or >41 weeks (N=482,706; 11.59%), (v) congenital anomalies (N=51,120; 1.23%), (vi) non-cephalic presentation (N=145,044; 3.48%), as well as births with missing data on these variables (N=96,117; 2.31%) (Fig S2). The final dataset included 3,168,273 low-risk births.

The APGAR score is a routine, standardized index conducted by a birth attendant at 1- and 5-minutes after birth. Newborns are scored 0-2 on five criteria (colour/peripheral cyanosis, heart rate, response to stimulation, muscle tone, and breathing), yielding a total score between 0 and 10 (6). Our primary outcome was a low-risk live-birth of a newborn with an APGAR score of 7 and below (≤7) at 5-minutes of life (hereafter, low APGAR-5’). Secondary outcomes were subcategories of low APGAR-5’: 0-2, 3-5 and 6-7, in line with Brazilian reporting standards (26).

#### Exposure assignment

Hourly 2-metre air and dew point temperatures (downloaded in Kelvin and converted to Celsius) were obtained for São Paulo state, at a 0.1° x 0.1° (∼9 x 9 km) spatial resolution, from the Copernicus Climate Data Store as part of the European Centre for Medium-Range Weather Forecasts’ (ECMWF’s) ERA5-Land reanalysis data (29). Municipality-level daily mean temperatures (°C) were calculated from hourly air temperatures arithmetically averaged by date over grid cells within municipality boundaries. Relative humidity (%) was calculated as the ratio between real vapor pressure and saturation vapor pressure obtained using Tetens equation with 2-meter dewpoint and air temperatures respectively.

Daily mean temperature was considered the best approximation of daily temperature exposures (accounting for both daytime and night-time temperatures). Our window of exposure was the day of delivery (lag 0) and day preceding delivery (lag 1), approximating childbirth and labour. Exposures were linked to outcomes by date and municipality of delivery. Population-weighted percentiles of daily mean temperature were calculated for São Paulo state over the study period (2013–2019). High and low temperatures were defined as the 95th and 5th percentiles of daily mean temperature, respectively.

#### Municipality-level metrics

Municipality-level deprivation data were obtained from Oswaldo Cruz Foundation’s Centre for Data and Knowledge Integration for Health (*Centro de Integração de Dados e Conhecimentos para Saúde*) (30). These data reflect the Brazilian Deprivation Index (*Índice Brasileiro de Privação*, IBP); a composite index based on the 2010 census data on low-income households (per capita income ≤50% minimal wage), illiterate individuals aged ≥7 years, and individuals with inadequate access to water and sanitation. We used population-weighted quintiles, based on deprivation at the countrywide level, and ordered from the least (1st quintile) to the most deprived (5th quintile) municipalities (Fig S1).

The Köppen classification system assigns climatic regions using temperature and precipitation and allows an understanding of regional biomes, weather and local infrastructural adaptation. Köppen designations for each municipality were obtained from Alvares *et al*. (25).

### Statistical analysis

Daily mean temperature and rate of low APGAR-5’ score (n cases / N low-risk births) in São Paulo state were plotted against day-of-year to highlight seasonal patterns. Scatterplots were used to examine the functional form of the unadjusted relationships between temperature, relative humidity and rate of low APGAR-5’ score at 0- and 1-day lags. The association between proposed effect modifiers and APGAR-5’ scores (≤7 vs ≥8) were assessed using Chi-square tests for independence.

We used a bi-directional time-stratified case-crossover design to estimate the short-term association between daily mean temperature and low APGAR-5’ score. This design has been widely used to assess acute impacts of environmental exposures on health, including birth outcomes (31,32). Cases act as their own controls, whereby exposure on the day of the health event (case day) is compared to exposure on otherwise similar days when the health event did not occur (control days). Control days were selected as the same day-of-week, calendar month and year as the case day, resulting in 3-4 control days per case. By design, this approach controls for day-of-week effects, seasonal and long-term trends, as well as time-fixed (or slow-changing) individual-level confounders (e.g. socio-economic position, age).

Conditional logistic regression was used to compare the likelihood of exposure to high and low temperature on case days versus matched control days. We applied distributed lag nonlinear models (DLNM) to capture potential nonlinear effects of temperature on the day of delivery and day prior (0-1 day lag). The temperature-outcome dimension was modelled using a natural cubic spline with one internal knot, while the lag dimension was modelled using a linear term. The number and positioning of spline knots were determined through minimisation of Akaike Information Criterion (AIC) and examination of diagnostic plots. Relative humidity (RH) was adjusted for as a time-varying confounder. RH was averaged over the 0-1 day lag and fitted using a spline with 3 degrees of freedom (*df*). The same model terms were used to analyse secondary outcomes. Effect estimates were expressed as the odds ratio (OR) of low APGAR-5’ score at the 95^th^ and 5^th^ percentiles of daily mean temperature, compared with exposure to the 50^th^ percentile. OR of low APGAR-5’ with exposures on 0 and 1 lag days before delivery (2-day cumulative) are presented, with reference to the contribution of individual lags where relevant.

#### Subgroup analysis

Subgroup analyses were used to assess potential effect modification by individual and area-level characteristics, including maternal age (<20 years, 20-34 years, >34 years), maternal race/ethnicity (Brown/*Parda*, Black/*Preta*, White/*Branca*, Asian-descent/*Amarela* and Indigenous*/Indígena*), maternal education (<12 years, ≥12 years), parity (nulliparous, primiparous, multiparous), timing of prenatal care initiation (in the first trimester of pregnancy, delayed), infant sex (male, female), municipality-level deprivation (in quintiles), and Köppen climate zones. Stratum-specific ORs were estimated using the same model terms as described above, to ensure comparability between groups. ORs for Köppen climate zones were centered at the population-weighted percentiles of daily mean temperature of either tropical or temperate regions.

#### Sensitivity analysis

As sensitivity analyses, models were respecified with (i) increased flexibility in the exposure-response curve (natural cubic spline with 2 internal knots); (ii) removal of humidity adjustment; and (iii) a different lag structure (lag 0 only; 0-6 day lag, fitted using a natural cubic spline with one internal knot in the lag and temperature dimensions). We also conducted an equivalent case-crossover analysis using conditional quasi-Poisson regression to ensure our findings were robust to adjustment for daily birth counts (rate denominator), overdispersion, and autocorrelation (33). Finally, to assess the potential selection bias caused by restricting our sample to low-risk births, we repeated the analysis on all singleton births between 2013-2019 in São Paulo state.

All analyses were conducted in R (4.2.3) using Rstudio and facilitated by the following packages: survival (34), dlnm (35) and splines (36). The study followed the STROBE reporting guidelines.

## Results

Between 2013 and 2019, 34,980 (1.1%) of the 3,168,273 newborns with low-risk births in São Paulo state had a low APGAR-5’ score (≤7). The annual rate peaked at 1.2% in 2015 and subsequently decreased to 1.0% by 2019 (Table S1). Low scores (≤7) were more common among male newborns and offspring of women who were younger (<20 years), nulliparous, had <12 years of education, self-identified as Brown/*Parda* or Black/*Preta*, and/or had later prenatal care initiation (Table 1).

**Table 1.** Characteristics of low-risk births (n=3,168,273) in São Paulo state, Brazil (2013-2019).

| Category | APGAR-5' score 0-7<br>N (%) | APGAR-5' score 8-10<br>N (%) | P-value* |
| --- | --- | --- | --- |
| **Totals** | 34,980 | 3,133,293 |  |
| **Maternal Age** |  |  |  |
| <20 years | 5,858 (16.8) | 404,187 (12.9) | $\chi^2 (2) = 454.82$,<br>$p < 0.001$ |
| 20-34 years | 23,764 (67.9) | 2,222,700 (70.9) |  |
| $\geq 35$ years | 5,358 (15.3) | 506,400 (16.2) | |
| Missing | 0 | 6 |  |
| **Maternal Education** |  |  |  |
| <12 years | 28,687 (82.2) | 2,353,730 (75.4) | $\chi^2 (1) = 878.15$,<br>$p < 0.001$ |
| $\geq 12$ years | 6,194 (17.8) | 768,931 (24.6) | |
| Missing | 99 | 10,632 |  |
| **Maternal Race/Ethnicity** |  |  |  |
| White/*Branca* | 17,909 (51.6) | 1,793,980 (57.7) | $\chi^2 (4) = 582.34$,<br>$p < 0.001$ |
| Brown/*Parda* | 14,171 (40.8) | 1,114,328 (35.8) |  |
| Black/*Preta* | 2,451 (7.1) | 178,441 (5.7) |  |
| Asian/*Amarela* | 152 (0.4) | 19,117 (0.6) |  |
| Indigenous/*Indígena* | 59 (0.2) | 4,607 (0.2) |  |
| Missing | 238 | 22,820 |  |
| **Parity** |  |  |  |
| Nulliparous | 19,247 (55.8) | 1,408,894 (45.8) | $\chi^2 (2) = 1391$,<br>$p < 0.001$ |
| Primiparous | 8,935 (25.9) | 1,007,129 (32.8) |  |
| Multiparous | 6,303 (18.3) | 659,598 (21.5) |  |
| Missing | 495 | 57,672 |  |
| **Prenatal Care Initiation** |  |  |  |
| During first trimester | 28,490 (84.7) | 2,627,835 (86.1) | $\chi^2 (1) = 54.208$,<br>$p < 0.001$ |
| After first trimester | 5,155 (15.3) | 425,084 (13.9) |  |
| Missing | 1,335 | 80,374 |  |
| **Newborn Sex** |  |  |  |
| Male | 19,996 (57.2) | 1,586,949 (50.7) | $\chi^2 (1) = 587.66,$<br>$p < 0.001$ |
| Female | 14,984 (42.8) | 1,546,344 (49.4) |  |
| **Brazilian Deprivation Index (IBP) \*\*** |  |  |  |
| 1 (Least deprived) | 11,659 (33.3) | 1,110,639 (35.5) | $\chi^2 (3) = 149.25,$<br>$p < 0.001$ ^ |
| 2 | 17,682 (50.6) | 1,580,246 (50.4) |  |
| 3 | 5,196 (14.9) | 403,342 (12.9) |  |
| 4 | 443 (1.3) | 39,060 (1.3) |  |
| 5 (Most deprived) | 0 (0.0) | 5 (0.0) |  |
| Missing | 0 | 1 |  |
| **Köppen Climate Zones** |  |  |  |
| Af - Tropical Rainforest (no dry season) | 2,171 (6.2) | 133,837 (4.3) | $\chi^2 (5) = 747.99,$<br>$p < 0.001$ |
| Aw - Tropical Savanna (dry winters) | 1,482 (4.2) | 201,809 (6.4) |  |
| Cfa - Temperate Humid-subtropical (no dry season, hot summer) | 6,971 (19.9) | 651,614 (20.8) |  |
| Cfb - Temperate Oceanic (no dry season, warm summer) | 20,744 (59.3) | 1,760,430 (56.2) |  |
| Cwa - Temperate Humid-subtropical (dry winter, hot summer) | 3,320 (9.5) | 351,980 (11.2) |  |
| Cwb - Temperate Highland-subtropical (dry winter, warm summer) | 292 (0.8) | 33,622 (1.1) |  |
| Missing | 0 | 1 |  |

\*P-values were obtained from the Chi-square test for independence. Missing values were excluded from the calculation.
^ IBP 5 was excluded from the Chi-square test due to small sample size.
\*\* The Brazilian Deprivation Index is based on the country as whole, allowing comparability of municipality-level deprivation across regions of Brazil.

Daily rates of low APGAR-5’ scores showed little seasonal variation (Fig 2). Based on population-weighted daily mean temperatures, the high (95^th^ percentile), moderate (50^th^ percentile), and low (5^th^ percentile) temperatures were 26.1°C, 20.9°C, and 14.8°C, respectively.

**Fig 2.**
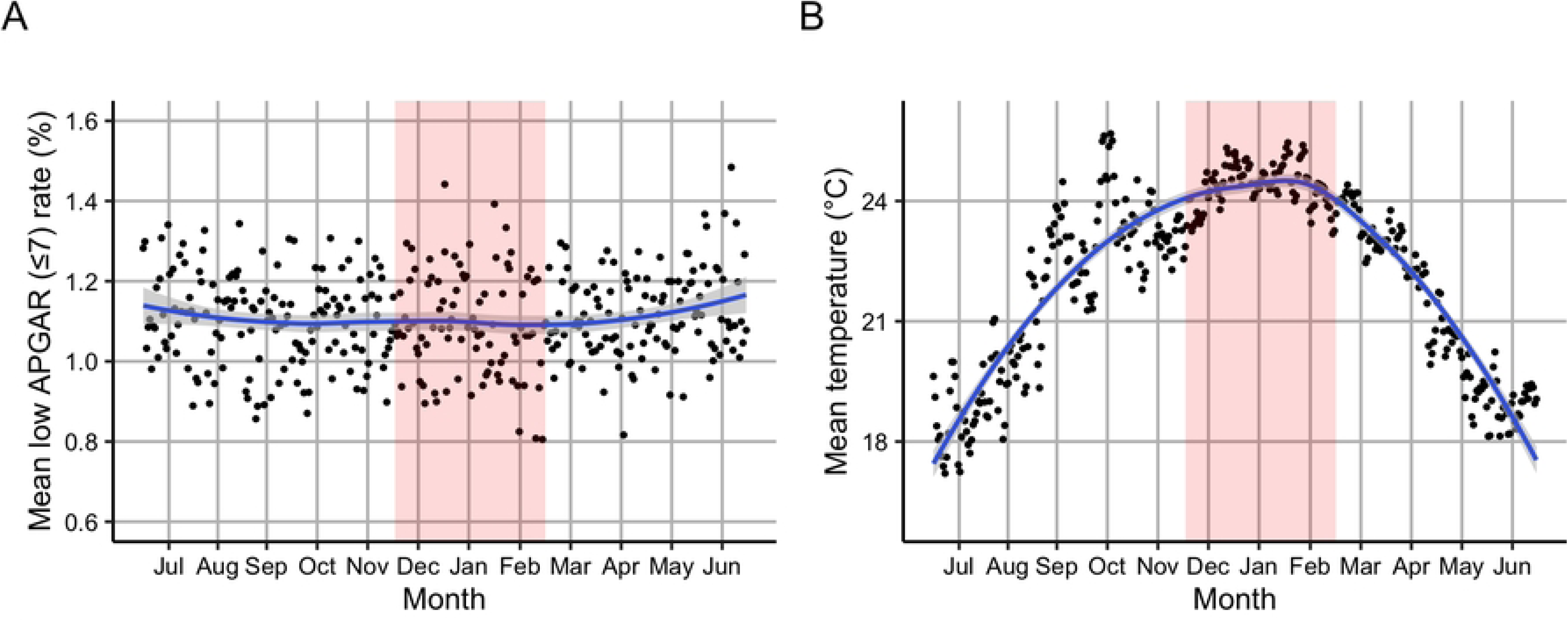
Seasonal patterns in daily mean low APGAR-5’ score (≤7) rate and daily mean temperature (°C). (A) Daily mean low APGAR-5’ (≤7) rate (%) among low-risk deliveries, and (B) daily mean temperature (°C), averaged (mean) by day-of-year across all municipalities in São Paulo state over the study period (2013–2019). The blue line was fitted using Locally Estimated Scatterplot Smoothing (LOESS) to illustrate seasonal trends. Summer months (December to February) are shaded in red.

Regression coefficients and fit statistics for models testing the association between daily mean temperature exposures and low APGAR-5’ score subcategories are presented in Table S2. Odds of low APGAR-5’ score (≤7) increased by 8% (OR: 1.08, 95% CI: 1.02-1.14) with exposure to high versus moderate temperature (26.1°C vs 20.9°C) 0-1 days before delivery (2-day cumulative) (Fig 3A). The effect of high temperature was largely confined to the day of delivery (lag 0; OR: 1.07, 1.00-1.15), with no evidence of association on the preceding day (lag 1; OR: 1.01, 0.94-1.07) (Table S3). There was no evidence that 2-day cumulative exposure to low temperature (14.8°C vs 20.9°C) was associated with low APGAR-5’ scores (≤7; OR: 0.95, 95% CI: 0.91-1.00).

**Fig 3.**
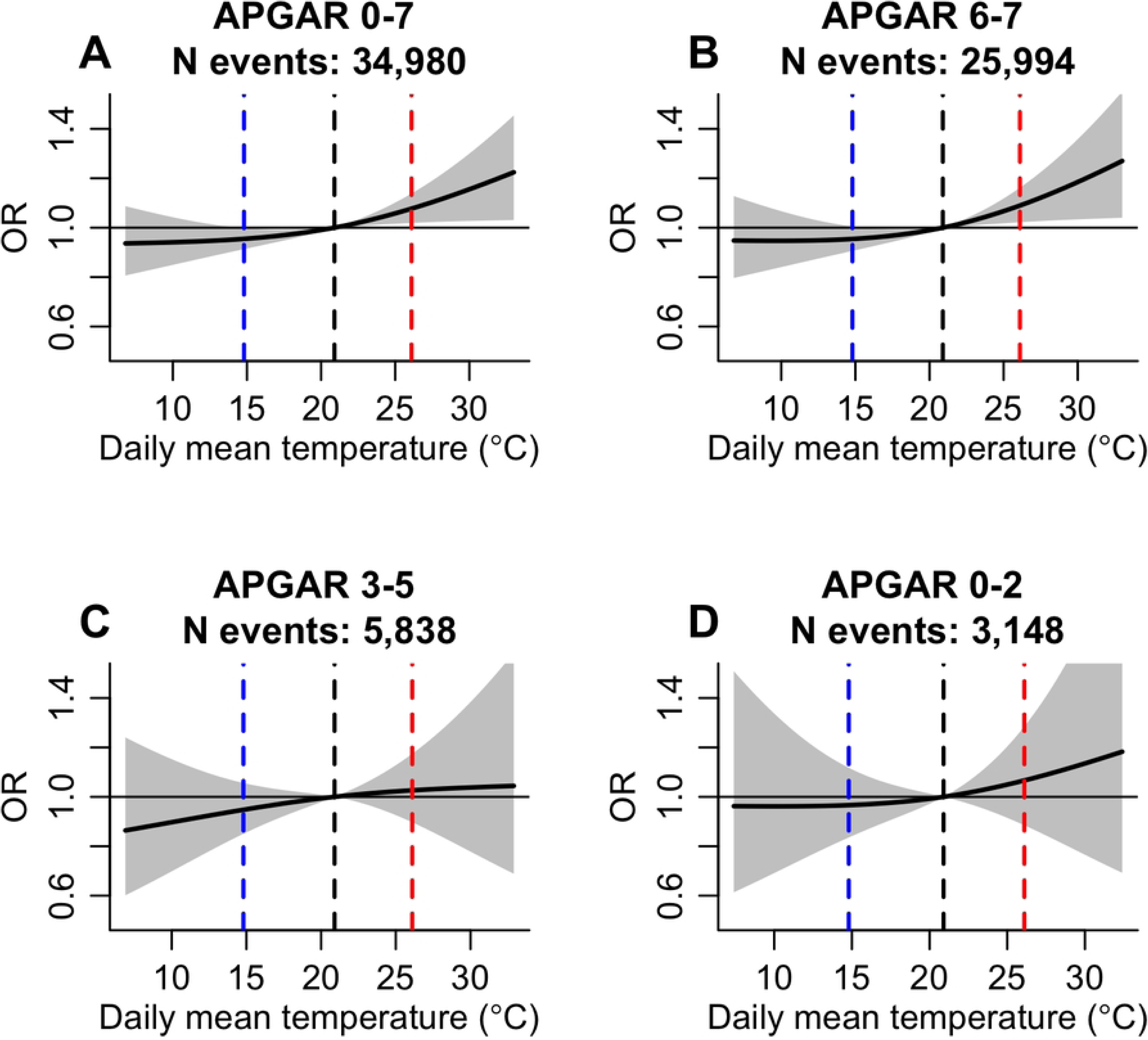
Association between ambient temperature exposures and odds of low-risk delivery with low APGAR-5’ score. Odds ratio (black line) and 95% confidence interval (grey shading) of low APGAR-5’ score (a); 0-7 (b) 6-7; (c) 3-5; and (d) 0-2 with exposure to daily mean temperatures 0-1 days before delivery (2-day cumulative). Predictions were centred at the 50th percentile of population-weighted daily mean temperature (20.9°C; black dashed line). Red dashed lines show the 95th percentile (26.1°C), and blue dashed lines show the 5th percentile (14.8°C) of population-weighted daily mean temperature.

A very similar exposure-response association was observed for low-risk deliveries with APGAR-5’ scores of 6-7 (Fig 3B) with odds increasing by 1.09 (95% CI: 1.02-1.16) with 2-day cumulative high temperature exposure (Table S3). Again, the effect of high temperature was strongest on the day of delivery (OR: 1.09, 1.01-1.18), with no evidence of a heat effect on the preceding day (OR: 1.00, 0.92-1.07). Odds of APGAR-5’ scores 3-5 or 0-2 did not change with 2-day cumulative temperature exposure (Fig 3C and 3D).

Fig 4 presents the 2-day cumulative OR of low APGAR-5’ score (≤7) with exposure to high (95th percentile) vs moderate (50^th^ percentile) temperature, stratified by characteristics of the population. We caution that the 95% confidence intervals for many OR estimates overlap with one, and very small sample sizes among Asian/*Amarela* and Indigenous subgroups hinder interpretation. There was a tendency towards an increased odds of low APGAR-5’ score with exposure to high temperature among all subgroups, except newborns of women with ≥12 years of schooling and newborns in the most deprived municipalities (4^th^ quintile). Among the most deprived municipalities (4^th^ quintile), 2-day cumulative low temperature exposure posed a greater risk (OR: 1.57, 95% CI: 1.09-2.25; Fig S3) than high temperature exposure (OR: 0.74, 95% CI: 0.44-1.24).

**Fig 4.**
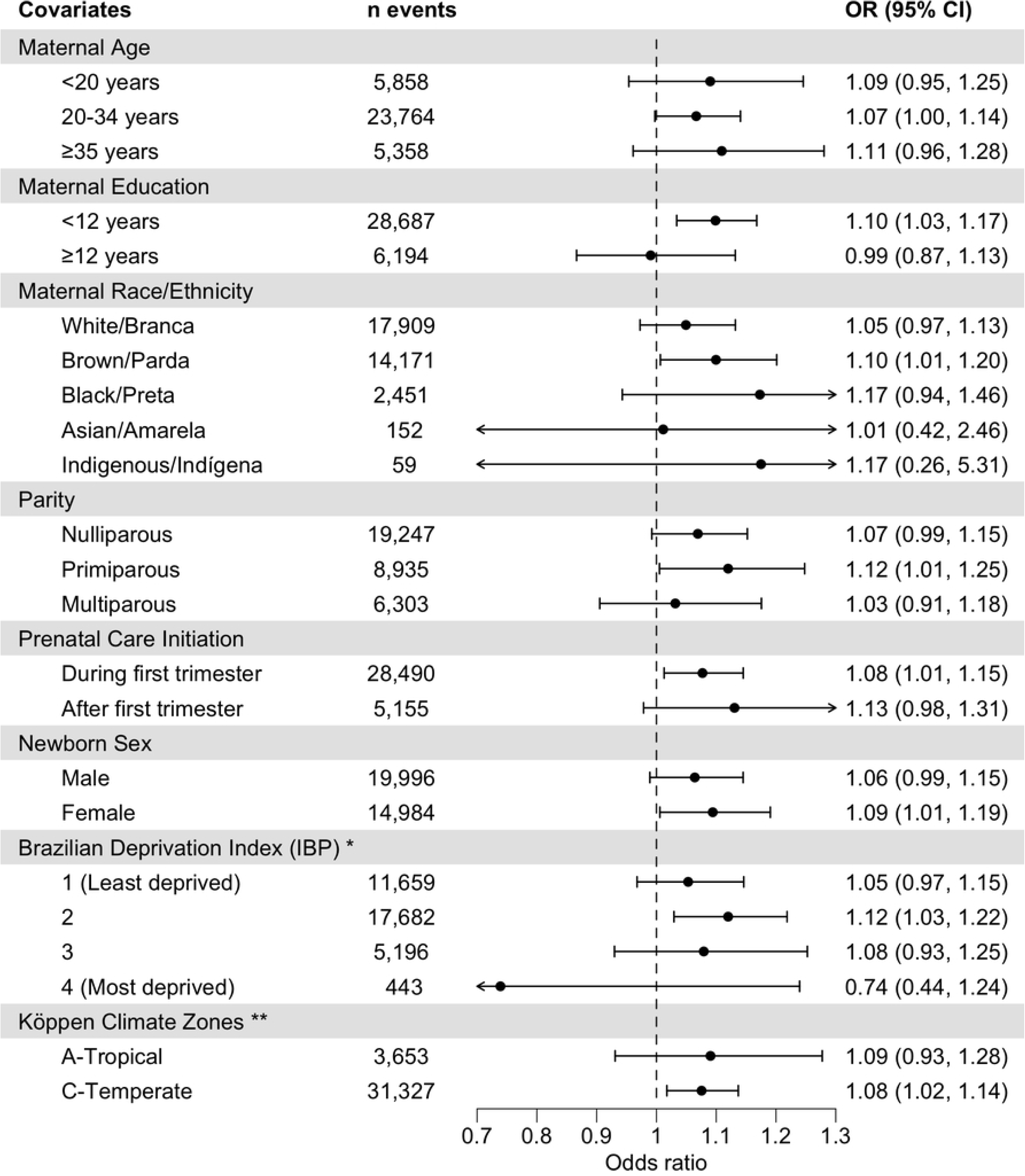
Effects of high temperature exposure on odds of low APGAR-5’ score (≤7), stratified by subgroup. Odds ratio (OR) and 95% CI of low APGAR-5’ score (≤7) at the 95th percentile (versus the 50^th^ percentile) of daily mean temperature 0-1 days before delivery (2-day cumulative). Temperature percentiles were calculated from population-weighted daily mean temperature in São Paulo state (2013-2019) and are 26.1°C (95^th^ percentile) and 20.9°C (50^th^ percentile) for all subgroups except Köppen climate zones, which were centered at respective population-weighted percentiles. Error bars represent a significant result at the 5% level where the 95% confidence interval does not include 1. * The Brazilian Deprivation Index is based on the country as whole, allowing comparability of municipality-level deprivation across Brazilian regions. No low-risk births with low APGAR-5’ (≤7) occurred in municipalities in the 5th deprivation quintile. ** For Tropical zones, the 50th percentile was 23.6°C and 95th percentile was 28°C. For Temperate zones, the 50th percentile was 20.6°C and the 95th percentile was 25.6°C.

Point estimates for the association between high temperature and low APGAR-5’ score (≤7) were similar across subgroups. We note minimal variation between strata of maternal age, newborn sex, and Köppen climate zones. However, differences are more apparent when Köppen climate zones are stratified further (Table S4); stronger heat-low APGAR-5’ associations are seen in Tropical Rainforest (Af, OR: 1.29, 95% CI: 1.04-1.62) and Temperate Humid-subtropical zones (Cfa, 1.15, 1.04-1.27). Parity and the timing of prenatal care initiation also exhibited very little variation across strata, although associations were slightly higher in primiparous women and those with delayed prenatal care initiation. Despite overlapping error bars between strata, more notable group differences were observed for maternal education, race/ethnicity and municipality-level deprivation (IBP), with stronger evidence of adverse effects observed in women with <12 years of schooling, Brown/*Parda* women, and municipalities in the 2^nd^ deprivation quintile. Finally, across subgroups, high temperature on the day of delivery (lag 0) generally posed the greatest risk (Table S5).

Effect estimates were robust to sensitivity analyses. Importantly, the stronger heat effect on lag 0 compared to lag 1 remained robust to any changes to model terms, lag structure, method or sample (Fig S4). The strength of evidence for an association between high temperature and low APGAR-5’ (≤7) increased with additional flexibility in the exposure-response relationship (Table S6, mod 4). Effect estimates were slightly lower when using the conditional quasi-Poisson model (adjusted for rate denominator, over-dispersion, and autocorrelation) (Table S6, mod 6) and when including higher-risk births (less restricted sample; see Table S7); however, the primary association was consistently driven by the moderately low APGAR-5’ subcategory (6-7; Table S7).

## Discussion

This study evaluated the impact of short-term ambient heat exposure on low APGAR-5’ scores among low-risk births in São Paulo state, Brazil. High temperatures on the day of and day preceding delivery was associated with 8% increased odds of low APGAR-5’ score (≤7), primarily attributable to exposure on the day of delivery (lag 0) and driven by newborns with APGAR-5’ scores of 6-7. Elevated risks were observed among infants born to primiparous women, with fewer than 12 years of schooling, who self-declared as Brown/*Parda*, and newborns delivered in municipalities in the second deprivation quintile.

Our findings are consistent with previous studies conducted across diverse geographic and climatic regions, which generally report an increased risk of low APGAR score with exposure to high temperature in late pregnancy. Exposure to hot days during the third trimester was found to decrease healthy APGAR-5’ scores (≥8) in rural Colombia by 0.6% (10), and overall score by 0.08 points in Australia’s Northern territory (12). Only one previous study, based in the Ningxia Hui region of Northwest China, examined the short-term effects of ambient temperature near the time of delivery. They found that same-day (lag 0) heat exposure was associated with 30.7% increased odds of very low APGAR-5’ (≤ 3) and, similar to our findings, a non-significant association was observed for exposure on the day before delivery (13). The lower effect estimate in our study may be explained by differences in climate zones. For instance, one study (37) found greater risks of heat-related preterm birth in arid-desert-cold climates (characteristic of Ningxia Hui) compared to humid-subtropical climates (characteristic of São Paulo). This suggests that populations in São Paulo might be better physiologically acclimatized or culturally adapted to heat than those in Ningxia Hui.

Another possibility is that our restricted ‘low-risk’ sample introduced selection bias, potentially underestimating the effects of heat by excluding the most vulnerable newborns. However, this is unlikely as the inclusion of higher-risk births in our sensitivity analysis actually yielded a smaller observed effect of ambient heat on newborns’ immediate health. Higher-risk births are typically admitted earlier and managed more intensively in specialised or tertiary care facilities equipped with air conditioning, where immediate interventions may mitigate the effects of ambient heat exposure. This may also explain the null association observed for newborns with APGAR-5’ scores of 0-5. Although we restricted our sample based on gestational age, birthweight, congenital anomalies and birth presentation, certain unobservable delivery risks – such as those related to maternal conditions or comorbidities – could not be excluded. Alternatively, the higher inherent baseline risks among these births might dilute the relative association with heat exposure. Notwithstanding, further research is needed to assess any mitigating effects of air conditioning on low APGAR-5’ score.

Our subgroup analysis supports previous heat-related perinatal health research in Brazil, which indicate inequities in the vulnerability to environmental stressors (22,23). Social determinants of health, like poverty and structural racism, may influence the exacerbating factors highlighted in the pathway between heat and low APGAR-5’ (Fig 1). Heat exposure is likely higher among lower income, less educated groups who may have more poorly insulated housing and lack air conditioning (38). Lower socio-economic position is associated with limited healthcare access (27), which may reduce opportunities for heat-related health promotion and/or delay diagnosis/treatment for obstetric conditions among groups with already reduced baseline health (39,40). However, we find only suggestive evidence of increased risks due to limited healthcare access, using delayed initiation of prenatal care as a proxy. Lower quality of care may also explain the observed association. Indeed, reduced referrals to specialised hospitals have previously been reported for people with lower education and from racially minoritised Black/*Preta* and Brown/*Parda* groups (28,41). Further research into the causes of differential vulnerability and targeted health promotion efforts are critical for maternal and newborn health equity.

Although there is a suggestion of a slight protective effect of low temperature on APGAR-5’ scores, as indicated in Colombia (10), the overall association was ultimately non-significant, consistent with studies in Northwest China and Australia’s Northern territory (12,13). However, low temperature was associated with an increased risk of low APGAR-5’ among newborns living in the most deprived municipalities, suggesting that exposure to cooler temperatures (as with warmer temperatures) interacts with social determinants to influence newborns’ immediate health. While not the primary focus of this study, due to the different biological mechanisms through which high and low temperatures may impact APGAR-5’ scores, we urge further investigation into differential cold vulnerability in this region.

Our study has several strengths. We conducted a hypothesis-driven study informed by research on pathways through which – and critical periods during which – heat exposure might impact low APGAR-5’ score. The case-crossover design was developed to study rare events caused by acute exposures (42) and findings were verified using extensive sensitivity analysis. Further research is needed to validate results in other Brazilian regions and contexts. However, the extensive coverage of SINASC data in São Paulo (99% of registered births) (43) and the minimal proportion (<0.5%) of births excluded due to missing APGAR-5’ scores, allow generalisation to low-risk births in Brazil’s most populous state (24).

We also acknowledge some key limitations. First, the case-crossover design assumes a constant rate of baseline outcome occurrence when not accounting for the exposure of interest (44). However, high temperature has been found to increase the birth-rate on the day of, and in the days following, exposure (45). This increase in births expands the at-risk population and, consequently, the number of deliveries with low APGAR-5’ scores. The estimated OR may thus capture the joint effect of temperature on both inducing labour (resulting in a low-risk birth) and lowering the newborn’s APGAR-5’ score. However, our sensitivity analysis using conditional quasi-Poisson regression – which is equivalent to the case-crossover design with adjustment for varying rate denominators (33) – also revealed increased risk of low APGAR-5’ (≤7) associated with high temperature on the day of delivery. This finding supports the robustness of our primary results.

Second, temperature exposures were constructed and assigned at municipality-level, which may not capture finer spatial variability, especially in larger municipalities where urbanisation and environmental conditions can vary significantly. However, we expect that the day-to-day fluctuations in temperature, which are essential for the present analysis, were still captured even if absolute temperature values are measured at a broader municipality-level. Additionally, some inconsistencies between weather station and ERA5-Land reanalysis data have been reported across Brazil, however daily temperature values from both sources are very strongly correlated (*r*=0.95) in São Paulo (46). Unavoidable exposure misclassification might also arise from the lack of data on air conditioning in both homes and hospitals. The vast proportion of births occur in health facilities, which typically have central air conditioning systems (26,47), however we are unable to ascertain the time of arrival at the facility or the presence of air conditioning in labour wards.

Third, caution is needed in interpreting the APGAR score as the predictive value of traditionally utilised cut-offs (<7 and <4) for long-term health outcomes has been challenged (48). However, previous research shows that even moderately low scores (e.g. 7) are associated with increased risks of neonatal morbidity and mortality (49). Herein, the outcome served to understand how ambient heat exposure during labour and delivery impacts the immediate health of the newborn. We defined low APGAR-5’ scores using both subcategories (0-2, 3-5, 6-7) and a threshold (≤7) aligned with Brazilian reporting conventions. Of note, APGAR-5’ scores were found to be less accurate for ascertaining the immediate health status of Black infants in the US, potentially due to the reliance on skin colour to ascertain oxygenation (50). Given the potential for similar misclassification in Brazil, APGAR-5’ scores may not always be appropriate when distinguishing between risk profiles of different ethnoracial groups. However, ambient temperature was not expected to alter the proportion of scoring mistakes by skin colour, therefore the higher observed heat-related risk among Brown/*Parda* populations is feasibly indicative of greater heat vulnerability.

## Conclusions

This study makes a valuable contribution to the Brazilian and global evidence base on the effects of ambient heat exposure on neonatal health. Findings suggest that acute heat exposure in low-risk births is a cause for concern, especially in light of rising temperatures in highly urbanised, temperate regions like São Paulo state (21). Our finding of increased heat-associated socio-economic vulnerability substantiates warnings that global warming will amplify the inequities in maternal and newborn health. Further research is needed to ascertain optimal ambient temperatures during labour and delivery and mediating bio-social pathways (e.g. delivery mode, care accessibility). However, we reinforce the urgent need for policies aimed at mitigating high temperature exposure near the time of delivery, including targeted health promotion initiatives and health infrastructure that is adapted at all levels.

## Data Availability

All data are publicly available in our GitHub repository at https://github.com/Michelle-Del-Carretto/LSHTM_HEAT_APGAR_CIDACS-CLIMA and can also be accessed using the DOI: https://doi.org/10.5281/zenodo.15090271. The repository contains the R scripts used for analysis and the datasets required to reproduce the results.

## Acknowledgments

The authors wish to express their gratitude to Huiqi Chen from the Faculty of Public Health and Policy at LSHTM for her invaluable support with the Quasi-Poisson analysis and Maxine Pepper from the Faculty of Epidemiology and Population Health at LSHTM for her important contribution regarding the conceptualisation of the joint outcome of delivery with low APGAR.

## Supporting information captions

**Fig S1. Geographic distribution of Brazilian Deprivation Index & Köppen climate zones in São Paulo state.**

**Fig S2. Flowchart of study inclusion and exclusion criteria.**

**Table S1. Total number of low-risk births by APGAR-5’ subcategory and year in São Paulo state (2013-2019).**

**Table S2. Regression coefficients and fit statistics for models testing the association between temperature and low APGAR-5’ subcategories.** Regression coefficients (Coef) are provided with their standard errors (SE), odds ratio (OR) and 95% confidence intervals (95% CI), p-values, and fit statistics. cb refers to the crossbasis object modelling the exposure-lag-response association. RH refers to relative humidity and was adjusted for as a confounder.

*Akaike Information Criterion

**Likelihood Ratio Test

**Table S3. Association between high daily mean temperature and low-risk birth with low APGAR-5’ score, by subcategory.** Odds ratio (OR) and 95% CI of low APGAR-5’ score subcategories (≤7, 6-7, 3-5, 0-2) with exposure to high temperature (95^th^ percentile, 26.1°C) relative to moderate temperature (50^th^ percentile, 20.9°C) 0-1 days before delivery (Lags 0-1; 2-day cumulative), on the day of delivery (Lag 0), and the day before delivery (Lag 1). Temperature percentiles were calculated from population-weighted daily mean temperature in São Paulo state (2013-2019).

**Fig S3. Effects of low temperature exposure on odds of low APGAR-5’ score (≤7), stratified by subgroup.** Odds ratio (OR) and 95% CI of low APGAR score (≤7) at the 5th percentile (versus the 50^th^ percentile) of daily mean temperature 0-1 days before delivery (2-day cumulative). Temperature percentiles were calculated from population-weighted daily mean temperature in São Paulo state (2013-2019) and are 14.8°C (5^th^ percentile) and 20.9°C (50^th^ percentile) for all subgroups except Köppen climate zones, which were centered at respective population-weighted percentiles.

* The Brazilian Deprivation Index is based on the country as whole, allowing comparability of municipality-level deprivation across Brazilian regions. No low-risk births with low APGAR-5’ (≤7) occurred in municipalities in the 5th deprivation quintile.

** Predictions for Köppen climate zones were centered at respective population-weighted percentiles. For Tropical zones, the 50th percentile was 23.6°C and 5th percentile was 17.4°C. For Temperate zones, the 50th percentile was 20.6°C and the 5th percentile was 14.6°C.

**Table S4. Association between high temperature and low APGAR-5’ (≤7), stratified by subcategories of Köppen climate zone.** Odds ratio (OR) and 95% CI of low APGAR-5’ score (≤7) with exposure to high daily mean temperature (95th percentile), relative to moderate temperatures (50th percentile), 0-1 days before delivery (Lags 0-1; 2-day cumulative), on the day of delivery (Lag 0), and the day before delivery (Lag 1). For Tropical zones, the 50th percentile was 23.6°C and the 95th percentile was 28°C. For Temperate zones, the 50th percentile was 20.6°C and the 95th percentile was 25.6°C.

**Table S5. Association between high temperature and low APGAR-5’ (≤7) score, stratified by subgroup.** Odds ratio (OR) and 95% CI of low APGAR-5’ score (≤7) with exposure to high daily mean temperature (95th percentile, 26.1°C), relative to moderate temperatures (50th percentile, 20.9°C) on the day of delivery (Lag 0) and the day before delivery (Lag 1). Percentiles were calculated from population-weighted daily mean temperature in São Paulo state (2013-2019). Predictions for Köppen climate zones were centred at respective population-weighted percentiles.

* The Brazilian Deprivation Index is based on the country as whole, allowing comparability of municipality-level deprivation across Brazilian regions. No low-risk births with low APGAR-5’ (≤7) occurred in municipalities in the 5th deprivation quintile.

** For Tropical zones, the 50th percentile was 23.6°C and 95th percentile was 28°C. For Temperate zones, the 50th percentile was 20.6°C and the 95th percentile was 25.6°C.

**Fig S4. Sensitivity analyses: Association between high temperature (lags 0 and 1) and low APGAR-5’ score (≤7).** Odds ratio (OR) of low APGAR-5’ score (≤7) with exposure to high versus moderate (95^th^ vs 50^th^ percentile, 26.1°C vs 20.9°C) daily mean temperatures on the day of delivery (lag 0) and day before delivery (lag 1) for models tested in sensitivity analyses. Percentiles were calculated from population-weighted daily mean temperature in São Paulo state (2013-2019). All models, unless specified otherwise, were performed on a restricted dataset of low-risk births.

Models are named following the convention: Lag structure – Crossbasis – Humidity adjustment

Crossbasis (CB) denotes the model terms used in the temperature dimension, then in the lag dimension. “lin” refers to linear. “ns” refers to a natural cubic spline, followed by the number of internal knots. The same convention applies to humidity. Humidity was always averaged (mean) over the included lag period. Where humidity was adjusted for, it is done so with using a natural cubic spline with 2 knots. ‘Hnone’ refers to no humidity adjustment.

For example, ‘Lag0-6 – CBns1-ns1 – Hns2’ refers to a regression analysis that modelled temperature exposures over 0-6 day lag, using a natural cubic spline with 1 internal knot in both temperature and lag dimensions, and adjusted for relative humidity using a natural cubic spline with 2 knots.

We conducted the Quasi-Poisson analysis excluding 0 strata (i.e. matched day-month-year-municipality sets without any low APGAR cases). Autocorrelation was adjusted for at lags 11 and 14.

*Performed on a larger sample of all singleton births between 2013-2019 in São Paulo state.

**Table S6. Sensitivity analyses: Association between high temperature (lags 0 and 1) and low APGAR-5’ score (≤7).** Odds ratio (OR) of low APGAR-5’ score (≤7) with exposure to high versus moderate (95^th^ vs 50^th^ percentile, 26.1°C vs 20.9°C) daily mean temperatures, 0-1 days before delivery (Lags 0-1; 2-day cumulative), on the day of delivery (Lag 0), and the day before delivery (Lag 1) for models tested in sensitivity analyses. Percentiles were calculated from population-weighted daily mean temperature in São Paulo state (2013-2019). All models, unless specified otherwise, were performed on a restricted dataset of low-risk births.

Models are named following the convention: Lag structure – Crossbasis – Humidity adjustment

Crossbasis (CB) denotes the model terms used in the temperature dimension, then in the lag dimension. “lin” refers to linear. “ns” refers to a natural cubic spline, followed by the number of internal knots. The same convention applies to humidity. Humidity was always averaged (mean) over the included lag period. Where humidity was adjusted for, it is done so with using a natural cubic spline with 2 knots. ‘Hnone’ refers to no humidity adjustment.

For example, ‘Lag0-6 – CBns1-ns1 – Hns2’ refers to a regression analysis that modelled temperature exposures over 0-6 day lag, using a natural cubic spline with 1 internal knot in both temperature and lag dimensions, and adjusted for relative humidity using a natural cubic spline with 2 knots.

We conducted the Quasi-Poisson analysis excluding 0 strata (i.e. matched day-month-year-municipality sets without any low APGAR cases). Autocorrelation was adjusted for at lags 11 and 14.

**Table S7. Sensitivity analysis: Association between high temperature and low APGAR-5’ in a less restricted sample.** Odds ratio (OR) of low APGAR-5’ score (≤7) with exposure to high versus moderate (95^th^ vs 50^th^ percentile, 26.1°C vs 20.9°C) daily mean temperatures, 0-1 days before delivery (Lags 0-1; 2-day cumulative), on the day of delivery (Lag 0), and the day before delivery (Lag 1). Temperature percentiles were calculated from population-weighted daily mean temperature. The sample included all singleton births between 2013-2019 in São Paulo state.

